# Demographic and socioeconomic factors predict maternal postpartum rehospitalization: a retrospective nuMoM2b dataset study

**DOI:** 10.1101/2022.02.23.22271437

**Authors:** Colin Wakefield, Martin G Frasch

**Affiliations:** Drexel University College of Medicine, Philadelphia, PA, USA; Health Stream Analytics LLC, Seattle, WA, USA; Dept. of OBGYN, CHDD, University of Washington, Seattle, WA, USA

## Abstract

The advent of artificial intelligence/machine learning (AI/ML) in medicine has opened opportunities for harnessing the power of ML to predict patient outcomes based on diverse data contained in the electronic medical records (EMR). Taken by themselves singular features represented by such diverse data are often not clearly predictive but combined in an ML modeling framework such so-called weak learners have the power to yield highly predictive models of health outcomes. We hypothesized that sociodemographic and basic, easily obtainable health characteristics of pregnant mothers strongly predict the risk of rehospitalization. Using the nuMoM2b dataset from NICHD and reproducible, open access AI/ML techniques we tested and validated this hypothesis. Among a large (n=10,038) and geographically diverse cohort of nulliparous women with singleton gestations, the present findings show that while some of these characteristics highlight the known disparities in health outcomes (race/ethnicity), most of them are modifiable through social policies and health counseling. These include access to and support during education, comprehensive health insurance, alleviating coming out of poverty, smoking, and unhealthy lifestyle habits. Across the four ML models deployed, the results are mutually reinforcing yielding a clear cohesive picture. This solution can be used to develop a prospective risk prediction for this important health outcome in pregnant mothers. The strengths of the presented solution are its focus on preventable maternal morbidities and risk factors, demonstration of the impact on adversely affected populations, and reproducible step-by-step executable, documented open-access code with relatively low computational requirements. This enables easy adoption and further development on this and other similar datasets to study the contributions and predictive power of various factors to maternal morbidities.

## Introduction

The promise of machine learning (ML) to predict patient outcomes based on diverse data from electronic medical records (EMRs) is only beginning to be realized.^1,2^

Since the release of the “Nulliparous Pregnancy Outcomes Study: monitoring mothers-to-be (nuMoM2b)” dataset, 30 papers have been published. However, no reproducible code has been made available yet to the scientific community to facilitate further study of this important dataset. In addition to providing such code, using a series of ML approaches we are addressing a central question which has not yet been explored on this dataset: what are the key predictors of maternal rehospitalization, a surrogate of maternal postpartum morbidity?

We hypothesized that maternal rehospitalization can be predicted reliably from the sociodemographic characteristics alone.

## Methods

The nuMoM2b is a prospective cohort study in which 10,038 nulliparous women with singleton pregnancies were enrolled from geographically diverse hospitals affiliated with 8 clinical centers. The data can be obtained via the NICHD DASH portal (https://dash.nichd.nih.gov/study/226675).

Full details of the study protocol have been published previously.^3^ In brief, women were eligible for enrollment if they had a viable singleton gestation, had no previous pregnancy that lasted more than 20 weeks of gestation, and were between 6 weeks 0 days’ gestation and 13 weeks 6 days’ gestation at recruitment. A common protocol and manual of operations were used for all aspects of the study. Each site’s local governing Institutional Review Board approved the study and all women provided informed written consent prior to participation.

Participant data were collected by trained research personnel during three antepartum study visits. These visits were scheduled to occur between 6 weeks 0 days’ and 13 weeks 6 days’ gestation (Visit 1), 16 weeks 0 days’ and 21 weeks 6 days’ gestation (Visit 2), and 22 weeks 0 days’ and 29 weeks 6 days’ gestation (Visit 3). A woman’s self-identified race and ethnicity was categorized as non-Hispanic white, non-Hispanic black, Hispanic, Asian or “other”. At least 30 days after delivery, trained and certified chart abstractors reviewed the medical records of all participants and recorded final birth outcomes and any readmissions to the hospital (rehospitalization).

To test our hypothesis, we built a classification model based on the respective maternal morbidity factors (Table 1) and the re-hospitalization rate response variable.

**Table 1.**
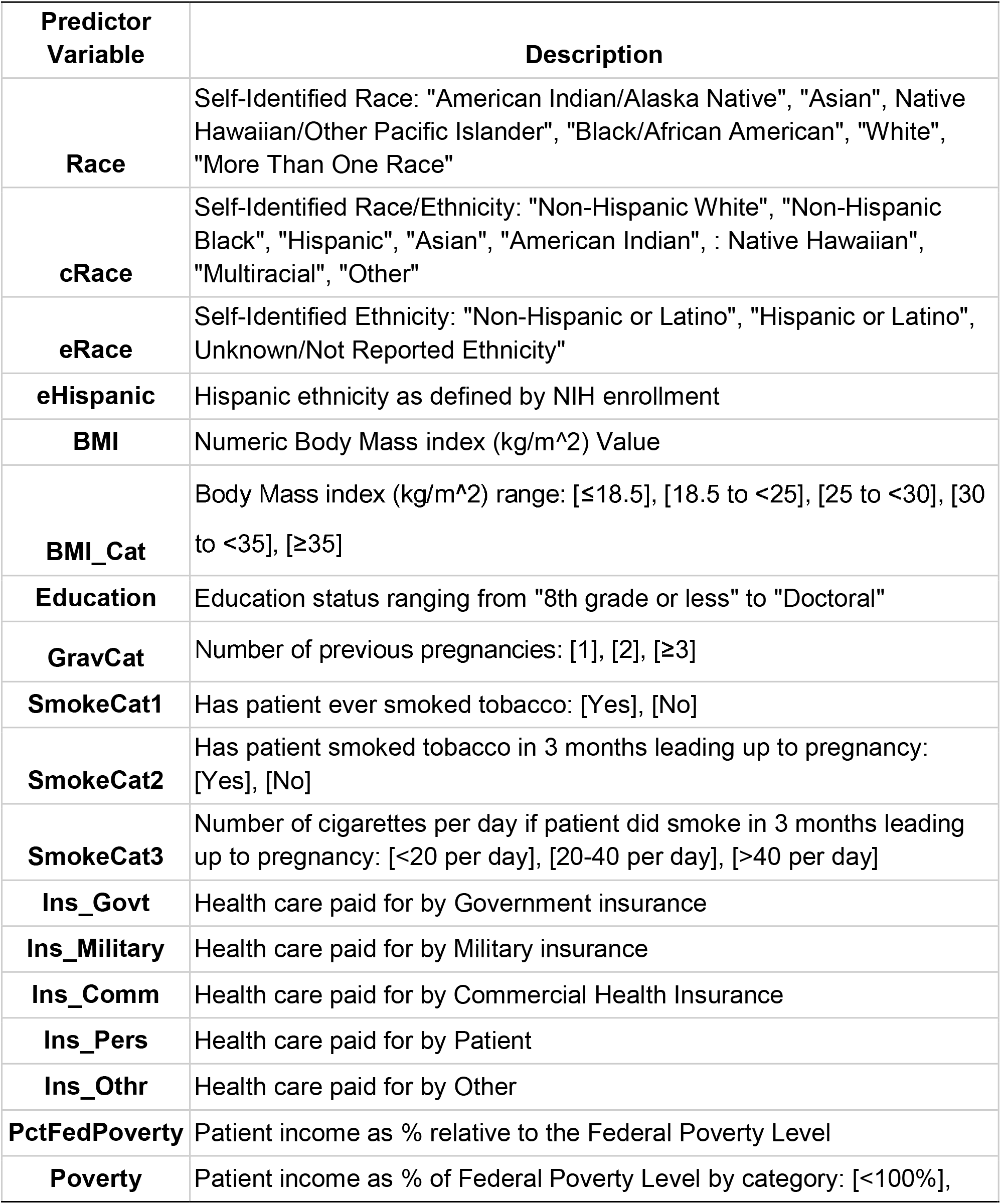
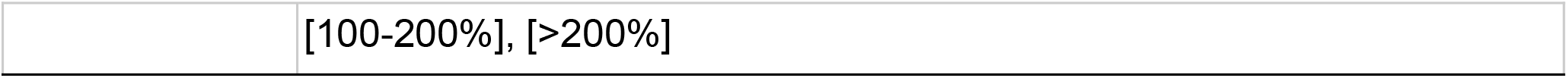
Description of predictor variables.

| **Predictor Variable** | **Description** |
| --- | --- |
| **Race** | Self-Identified Race: "American Indian/Alaska Native", "Asian", "Native Hawaiian/Other Pacific Islander", "Black/African American", "White", "More Than One Race" |
| **cRace** | Self-Identified Race/Ethnicity: "Non-Hispanic White", "Non-Hispanic Black", "Hispanic", "Asian", "American Indian", "Native Hawaiian", "Multiracial", "Other" |
| **eRace** | Self-Identified Ethnicity: "Non-Hispanic or Latino", "Hispanic or Latino", "Unknown/Not Reported Ethnicity" |
| **eHispanic** | Hispanic ethnicity as defined by NIH enrollment |
| **BMI** | Numeric Body Mass index (kg/m<sup>2</sup>) Value |
| **BMI_Cat** | Body Mass index (kg/m<sup>2</sup>) range: [$\leq 18.5$], [18.5 to $< 25$], [25 to $< 30$], [30 to $< 35$], [$\geq 35$] |
| **Education** | Education status ranging from "8th grade or less" to "Doctoral" |
| **GravCat** | Number of previous pregnancies: [1], [2], [$\geq 3$] |
| **SmokeCat1** | Has patient ever smoked tobacco: [Yes], [No] |
| **SmokeCat2** | Has patient smoked tobacco in 3 months leading up to pregnancy: [Yes], [No] |
| **SmokeCat3** | Number of cigarettes per day if patient did smoke in 3 months leading up to pregnancy: [$< 20$ per day], [20-40 per day], [$> 40$ per day] |
| **Ins_Govt** | Health care paid for by Government insurance |
| **Ins_Military** | Health care paid for by Military insurance |
| **Ins_Comm** | Health care paid for by Commercial Health Insurance |
| **Ins_Pers** | Health care paid for by Patient |
| **Ins_Othr** | Health care paid for by Other |
| **PctFedPoverty** | Patient income as % relative to the Federal Poverty Level |
| **Poverty** | Patient income as % of Federal Poverty Level by category: [$< 100\%$], |
|  | [100-200%], [>200%] |

The analyses have been performed in R 4.1.1 and h2o 3.34.0.3. Training and validation were done in 80:20 split with random 10-fold cross-validation. The code, results, and the <u>notebook</u> for validation and extension of the presented solution have been open-sourced.^6^ The following results are reported for the Distributed random forest ML model and were similar for Decision Tree, NaÏve Bayes Classifier and AutoML approaches, all reported in the shared notebook.

## Results

Of the 10,038 women enrolled in this prospective cohort, 9,470 were eligible for the present analysis (see Figure 1 in ^3^). Among eligible women, 5,721 (60.4%) were non-Hispanic white, 1,307 (13.8%) were non-Hispanic black, 1,586 (16.7%) were Hispanic, 379 (4.0%) were Asian, and 477 (5.0%) were of another race/ethnicity. For the outcome variable, there were a total of 8,776 participants’ data available for ML training to predict rehospitalization which was reported in 154 of the cases. To characterize model performance on this imbalanced dataset, we used precision/recall AUROC (prAUROC) which achieved a 0.98 predicting the outcome with an accuracy of 98.3% (CI 98.1%-98.4%) and an associated log loss of 0.14 (CI 0.12-0.19) (Fig. 1). The most important predictors were BMI, percentage of federal poverty level, education level, number of previous pregnancies, and race/ethnicity.

**Figure 1.**
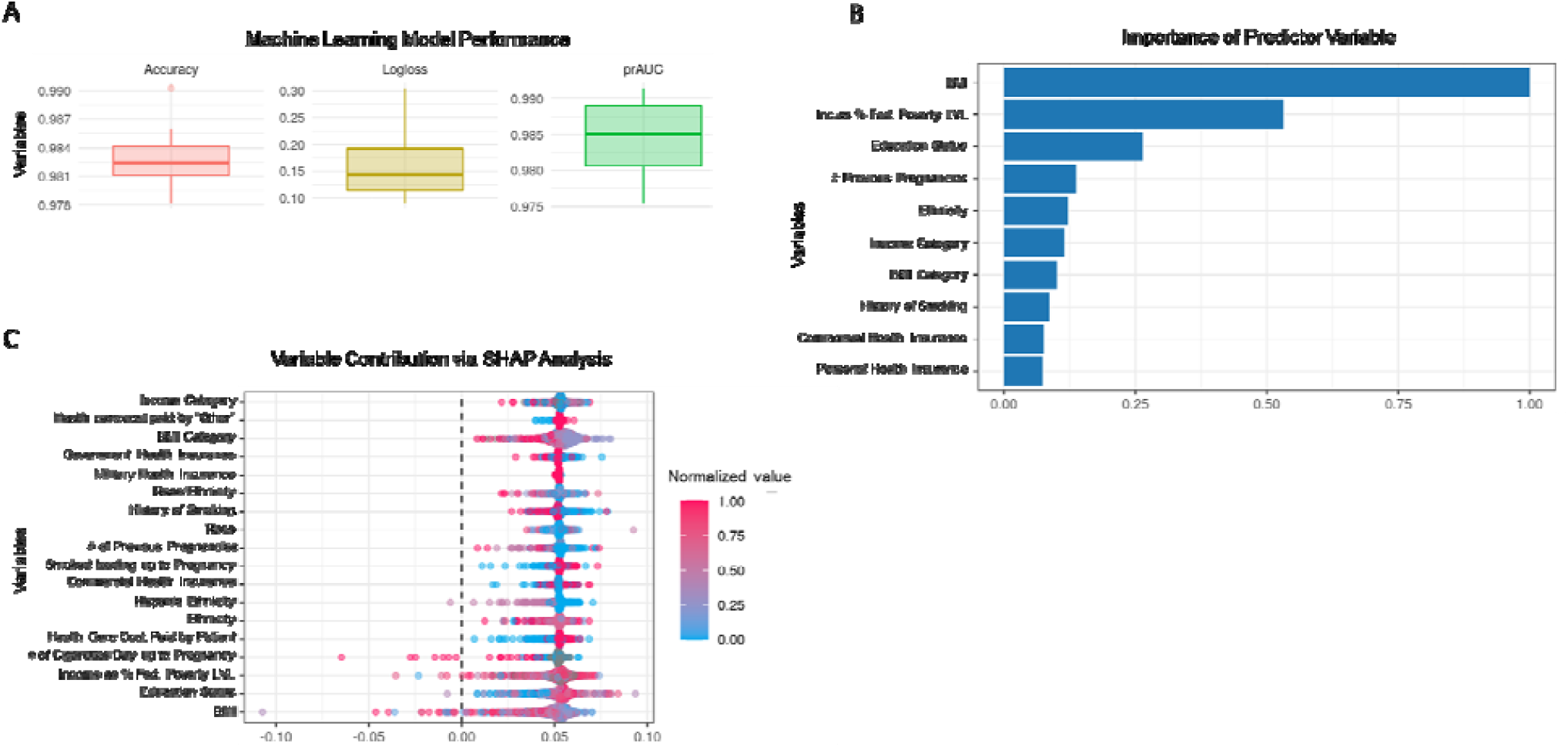
Re-admission to the hospital: main predictors and model performance. Distributed random forest machine learning (ML) model to predict rehospitalization. **A**. ML model performance: prAUC, precision AUC, an AUC metric better suited for this imbalanced dataset. **B**. Feature importance: contribution of the individual sociodemographic characteristics to the prediction. **C**. SHAP analysis: a different representation of the individual contributions of the considered predictor variables to the overall performance of the ML model.

## Discussion

The major contribution of the psychosocial determinants to health trajectories, especially during pregnancy, has been recognized,^5^ yet in the U.S. it has not been acted upon at the scale it warrants. Among a large and geographically diverse cohort of nulliparous women with singleton gestations, we show that while some of the predictors of rehospitalization highlight the known disparities in health outcomes (race/ethnicity), as reported in part for non-Hispanic black women^3^, most of the predictors are modifiable through social policies and health counseling. These include access to and support during education, comprehensive health insurance, alleviating coming out of poverty, smoking, and unhealthy lifestyle habits. Across the four ML models deployed, the results are mutually reinforcing yielding a clear cohesive picture.

Considering the recent <u>White House initiative</u> to reduce maternal morbidity and mortality, we hope these findings are especially timely on policy level and can be used to develop a prospective risk prediction for this and other important health outcomes of pregnancy.

## Data Availability

The code is available as follows:
https://doi.org/10.5281/zenodo.5785159
https://martinfrasch.github.io/maternal_nu_data.nb.html
The raw data can be obtained directly from NICHD via the NICHD DASH portal (https://dash.nichd.nih.gov/study/226675).

https://martinfrasch.github.io/maternal_nu_data.nb.html

## Notes

### Competing Interest Statement

MGF holds patents on fetal monitoring. MGF advises and holds stock in Delfina, a maternal-fetal health AI company.

### Clinical Protocols

https://dash.nichd.nih.gov/study/226675

### Funding Statement

This study did not receive any funding.

### Author Declarations

ClinicalTrials.gov Identifier: NCT01322529 IRB of Eunice Kennedy Shriver National Institute of Child Health and Human Development (NICHD) gave ethical approval for this work. The data was collected originally by NICHD Division/Branch/Center: DER - Pregnancy and Perinatology Branch (PPB) with NICHD Research Networks and Initiatives: Nulliparous Pregnancy Outcomes Study: Monitoring Mothers-to-Be (nuMoM2b) Study Description: NuMoM2b studied underlying, interrelated, mechanisms of adverse pregnancy outcomes (e.g., preterm birth, preeclampsia, fetal growth restriction) in pregnant women with no previous pregnancy lasting 20 weeks-0 days or more estimated gestational age (nulliparas). The network included 8 clinical sites, subsites, NICHD, and a data center. Women enrolled early in pregnancy and were followed through delivery, with study visits at 6 weeks-0 days to 13 weeks-6 days, 16 weeks-0 days to 21 weeks-6 days, 22 weeks-0 days to 29 weeks-6 days gestation, and delivery. Data collected through interviews, self-administered questionnaires, clinical measurements, ultrasounds, and medical records review included demographic, psychosocial, dietary, physiologic, health, and pregnancy outcome information. Subsets of participants enrolled in sleep disordered breathing, sleep patterns and quality, and fetal adrenal gland substudies. Source Repository: DASH https://dash.nichd.nih.gov/study/226675

